# Healthcare providers’ perceptions of barriers and facilitators to implementation of an acute traumatic brain injury clinical practice guideline: a CFIR-informed qualitative study at a regional referral hospital in Northern Tanzania

**DOI:** 10.64898/2026.10.04.26364725

**Authors:** Vivian F. Saria, Gregory G. Zaccheus, Joao Ricardo Nickenig Vissoci, Francis Sakita, Winfrida C. Mwita, Linda Minja, Timothy Antipas Peter, João Vítor Perez de Souza, Kim Madundo, Blandina T. Mmbaga, Catherine A. Staton, Anna Tupetz

## Abstract

**Background:** Traumatic brain injury (TBI) is a major cause of morbidity and mortality in low-and middle-income countries (LMICs), where the burden of injury is disproportionately high, and healthcare resources are often limited. Clinical Practice Guidelines (CPGs) improve the consistency and quality of TBI care by shifting medical decision-making from individual judgement to evidence-based recommendations. The strong link between improved TBI outcomes and CPG adoption has largely been established in high-income country (HIC) hospital settings; evidence on implementing TBI CPGs in LMIC hospital settings is limited.

**Objective:** This study explored healthcare providers’ experiences to identify barriers and facilitators to implementing a TBI CPG in the Emergency Department (EMD) at Kilimanjaro Christian Medical Center in Northern Tanzania, guided by the Consolidated Framework for Implementation Research 2.0 (CFIR).

**Methods:** We conducted semi-structured in-depth interviews (IDIs) with doctors and nurses who regularly managed TBI patients at the EMD. We purposively sampled participants between June and August 2023. The research team analysed data using thematic content analysis guided by CFIR domains, developing codes and memos iteratively.

**Results:** 7 nurses and 8 doctors with an average of 5 years of experience caring for patients in the EMD completed in-person, in-depth interviews. Most providers reported frequent exposure to TBI cases and regular use of the CPG in routine clinical practice. However, not all participants reported consistent CPG use, highlighting variation in implementation among providers. Key facilitators included strong interprofessional teamwork, readily accessible guideline materials in clinical areas, availability of diagnostic imaging, and perceived improvements in patient outcomes. Barriers to consistent adherence included ICU bed shortages, limited operating theatre capacity, equipment and medication stock-outs, high patient volumes, staffing pressures, and a lack of structured refresher training or formal audit and feedback mechanisms. Variability in procedural confidence, particularly among less experienced providers, also affected implementation consistency.

**Conclusion:** Our findings show that a TBI CPG is well accepted and largely integrated into routine care among experienced providers. However, sustainability depends on strengthening training, mentorship, audit, and feedback systems for CPG use and on addressing system-level resource constraints. Institutional investment in capacity building is essential to ensure consistent and sustained CPG uptake in our setting.

## INTRODUCTION

Traumatic brain injury (TBI) has the highest incidence of all common neurological disorders and poses a substantial public health burden (1,2). The burden of TBI injuries is disproportionately high in low-and middle-income countries (LMICs), where road traffic injuries, personal violence, and limited healthcare resources contribute substantially to its incidence and severity (3–5). Patients with TBI in LMICs have more than twice the risk of death following TBI compared to patients in high-income countries (HICs) (6–8). Although evidence-based clinical practice guidelines (CPGs) have been developed to improve acute TBI management and patient outcomes, their implementation remains inconsistent in many LMIC settings (9,10). In high-income-country hospital settings, improved TBI outcomes have been attributed to the widespread adoption of CPGs for acute TBI management (10). Although much of the evidence originates from high-income countries, structured TBI protocols have shown promise in improving care processes across diverse healthcare settings (1). CPGs promote standardized, evidence-based care and reduce unwarranted variation in TBI care (11,12). However, variations in guideline quality, scope, and contextual applicability may influence their interpretation, adaptation, and implementation, particularly in resource-limited environments (13,14). Understanding the contextual factors and healthcare providers’ experiences with CPGs that shape implementation is therefore critical, particularly in resource-constrained emergency care settings (15).

At the Kilimanjaro Christian Medical Center (KCMC) in Northern Tanzania, a prospectively collected TBI registry was established in 2013 to inform quality improvement efforts. This registry guided the development and adaptation of a TBI CPG in 2016, which has since been used across the Emergency Medicine Department (EMD) and by staff from General Surgery and Anesthesia. The CPG aims to harmonize acute TBI care, reduce variation in clinical decision-making, and ensure evidence-based management within a resource-constrained setting (10). This initiative is informed by data from the institutional TBI registry, which has previously described the patient population and injury characteristics and collected clinical data, highlighting opportunities for standardization and quality improvement (16).

This study aims to describe providers’ perceptions of barriers and facilitators of TBI CPG implementation, using the Consolidated Framework for Implementation Research (CFIR) constructs (17). By analysing interventional characteristics, inner-and outer-setting influences, individual providers’ factors, and implementation processes, this study identifies context-specific barriers and facilitators relevant to the sustained use of TBI CPGs in an LMIC emergency care setting.

## 2. MATERIALS AND METHODS

### 2.1. Study Design

This descriptive qualitative study focused on individual Healthcare Professionals’ (HCPs) experiences with the TBI CPG. The Kilimanjaro Christian Medical University College Ethics Committee approved the study under IRB protocol number 2545. All participants provided written informed consent. The study was reported in accordance with the Consolidated Criteria for Reporting Qualitative Research (COREQ) guidelines (18), (Supplemental Material 2), and the completed checklist is provided as supplementary Table 1.

### 2.2. Research Team and Reflexivity

The PI (VFS) and one Tanzanian research assistant (GG) conducted the interviews after receiving training in qualitative data collection methods from the research team. The PI is a registered female nurse with a Master of Science in Critical Care and Trauma, and the other data collector was a medical doctor. Both data collectors provided patient care at KCMC and might have been known to some participants. The PI served as a nurse leader in the Nursing Administration office but did not work in the EMD. The Tanzanian research assistant (GG) intermittently worked in the EMD and was familiar with the CPG.

### 2.3. Study Setting

This study took place in northern Tanzania. Tanzania is a lower-middle-income country in East Africa with an estimated population of 59 million. The Kilimanjaro Christian Medical Center (KCMC) is an urban tertiary-care referral hospital in the Kilimanjaro region of northeastern Tanzania, serving patients from both urban and rural areas. The hospital is the country’s third-largest facility, with 671 beds and a regional training center for various healthcare professions. The KCMC EMD treats approximately 2000 trauma cases annually, of which 1000, or roughly 6% of all emergency visits, are TBI cases (16, 19). KCMC offers inpatient and outpatient services across multiple specialties, providing follow-up and ongoing medical care after discharge. The hospital serves more than 60,000 patients yearly. The TBI mortality rate at KCMC is high, at 47% for severe (Glasgow Coma Score (GCS) <9) TBI patients and 13% for moderate (GCS 9-13) TBI patients (16). After initial CPG implementation in 2016, mortality decreased by an estimated 1.6% per month during the 9 months post-implementation (10). Patients with mild and moderate TBIs are usually admitted to general surgery departments equipped to manage their injuries. Severe TBI patients are admitted to the surgical intensive care unit (SICU), staffed by a few nurses specialised in critical care and trauma and equipped with ventilators and monitors. The EMD has access to basic laboratory and radiological services, including X-ray and computed tomography (CT).

During the study period, from June 2022 to August 2023, EMD staff included approximately 22 nurses, 4 emergency physicians, and 2 trauma nurse specialists. Clinicians from other departments who frequently evaluate TBI patients at the EMD include consultant neurosurgeons, anaesthetists, and general surgery residents.

### 2.4. Participant recruitment and sampling

Eligible participants included emergency medicine physicians, medical doctors, surgical residents, nurses (registered nurses and nurse specialists), anaesthesiologists, and general surgeons responsible for evaluating TBI patients at EMD. The nurse coordinator and the head of the EMD identified eligible HCPs through purposive sampling. Purposive sampling is the deliberate selection of an informant based on their potential to generate useful data for the study. (20). This sampling technique was chosen because it enabled the researcher to select participants with varying years of experience, exposure to the TBI CPG, and decision-making authority at KCMC, as well as professional roles in managing TBI patients, as this population was deemed to have rich information on the subject matter. We approached HCPs in person during their working hours in the EMD to introduce the study and invited them to participate at a convenient time outside of work hours. Participation was voluntary. HCPs who were not involved in the care of TBI patients or were not working at the EMD, as well as those with less than 1 year of professional experience in their current role, were excluded from this study. Participants signed informed consent forms before interviews began.

### 2.5. Data Collection

The PI developed an initial semi-structured interview guide informed by the Consolidated Framework for Implementation Research (CFIR) (17). The guide was organized around key CFIR domains, including intervention characteristics, inner setting, outer setting, and individual characteristics, to systematically explore multilevel providers’ perceptions of barriers, experiences, and facilitators to TBI CPG implementation. The research team developed the guide in English, translated it into Swahili with bilingual research assistants, back-translated it, and pilot-tested it. The interview guide underwent three revision cycles based on feedback from the rest of the research team using it (Supplemental material 1). VFS and GG pilot-tested the interview guide together, as they are both HCPs who would have met the study’s inclusion criteria. The pilot process allowed the researchers to resolve wording and structure issues and identify questions that were difficult to understand or required further probing. The researchers did not include the pilot interview data in the analysis. Demographic and professional characteristics were also collected (Supplementary material 1). We conducted all interviews in person after working hours in the hospital setting between June 2022 and August 2023. We conducted participant interviews in a quiet, private office at KCMC. Interviews lasted 30-45 minutes. The average length was 35 minutes. VFS or GG conducted the interviews, which were audio-recorded with only the participant present. All interviews were conducted in Swahili and transcribed verbatim to ensure consistency. Bilingual Tanzanian research assistants from VFS and GG then translated them into English. Although back-translation was not undertaken, the principal investigators, VFS, and GG ensured rigor by jointly cross-checking all English translations against the original Swahili transcripts to ensure accuracy and consistency. The data collectors, VFS and GG, received prior training in qualitative data collection and analysis from AT through a didactic workshop and practice session on coding. Throughout data collection and coding, the team met regularly to review transcripts, identify topic areas requiring further probing by the interviewers, and add follow-up questions as needed.

### 2.6. Data Coding and Analysis

#### Codebook development

The data collectors developed the first version of the codebook in October 2022 using an inductive, thematic coding approach. The rest of the research team then provided feedback. VFS and GG independently analysed the first three transcripts and developed English codes. We developed a preliminary codebook based on these transcripts. We then used it to guide coding of the remaining interviews, adding new codes and refining existing ones as needed throughout the analysis. We prepared a codebook and a participant summary in English to facilitate regular team discussions on emerging themes. We then deductively mapped emergent codes and themes to the five domains of the Consolidated Framework for Implementation Research (CFIR): Intervention Characteristics (e.g., perceptions of the TBI CPG’s complexity, adaptability, and relative advantage); Outer Setting (e.g., health system priorities, resource constraints, and external pressures); Inner Setting (e.g., EMD infrastructure, departmental culture, and communication networks); Characteristics of Individuals (e.g., providers’ knowledge, beliefs, and confidence in using the CPG); and Process (e.g., training, supervision, feedback, and monitoring of CPG implementation). This mapping provided a structured framework for identifying barriers and facilitators to the CPG implementation in the local context.

The team conducted coding in a Microsoft Excel spreadsheet to support collaboration and communication. The two data collectors, VFS and GG, double-coded the interviews. When they disagreed on a code, VFS and GG discussed it; if they could not reach consensus, a third reviewer (AT), experienced in qualitative analysis, made the final decision.

After one month of data collection, we performed a preliminary analysis of 4 transcripts to discuss the preliminary codebook and identify areas for improvement in the interview guide. Following this review, we made minor revisions to the interview guide to improve clarity and probing. In this process, we did not add any questions or topics. We repeated the analysis after every 4 interviews and, after 12 interviews, decided that data saturation had been reached. Data saturation is achieved when participant responses repeat information already obtained, and little new information is gained (21). We then purposely recruited additional EMD nurses to ensure a balance across professions, as well as HCPs who began working at the EMD after the CPG’s implementation in 2018. We conducted an additional 3 interviews with healthcare providers in August 2023. We did not repeat data collection, and we did not return transcripts to participants for comment or correction.

After 15 interviews, the full research team reviewed the final codebook and exemplary quotes for each theme and code. The team validated themes by sharing the themes and preliminary analysis documents to reach consensus. The team then further analysed and interpreted the data by creating descriptive content memos.

## RESULTS

We report findings from IDIs with 15 participants, exploring providers’ experiences with the TBI CPG in the Emergency Medicine Department (EMD), barriers and facilitators to its use, and perceived effects on providers’ and patients’ outcomes. Participants had a mean (SD) age of 32 (6.7) years, with an average of 5 (5.2) years of experience at KCMC and 4 (2.5) years in the EMD. Eight participants were male (53%). The study population included 8 doctors (53%) and 7 nurses, representing a range of speciality training and clinical experience (Table 1). Only two participants were working in the EMD during the initial implementation of the TBI CPG; the remaining participants joined the department after implementation and were introduced to the guideline during routine practice and orientation. Analysis of interview transcripts identified three overarching themes related to implementing the TBI CPG: healthcare providers’ experiences with the guideline, facilitators of its implementation, and barriers to consistent use (Figure 1). These themes reflect individual, organisational, and system-level factors that influence guideline uptake in the emergency department. The findings were presented in three themes: healthcare providers’ experiences, facilitators of CPG implementation, and barriers to CPG implementation. Following the inductive coding process, themes and subthemes were inductively mapped to the Consolidated Framework for Implementation Research (CFIR) domains to facilitate the interpretation of implementation (Table 2).

**Figure 1.**
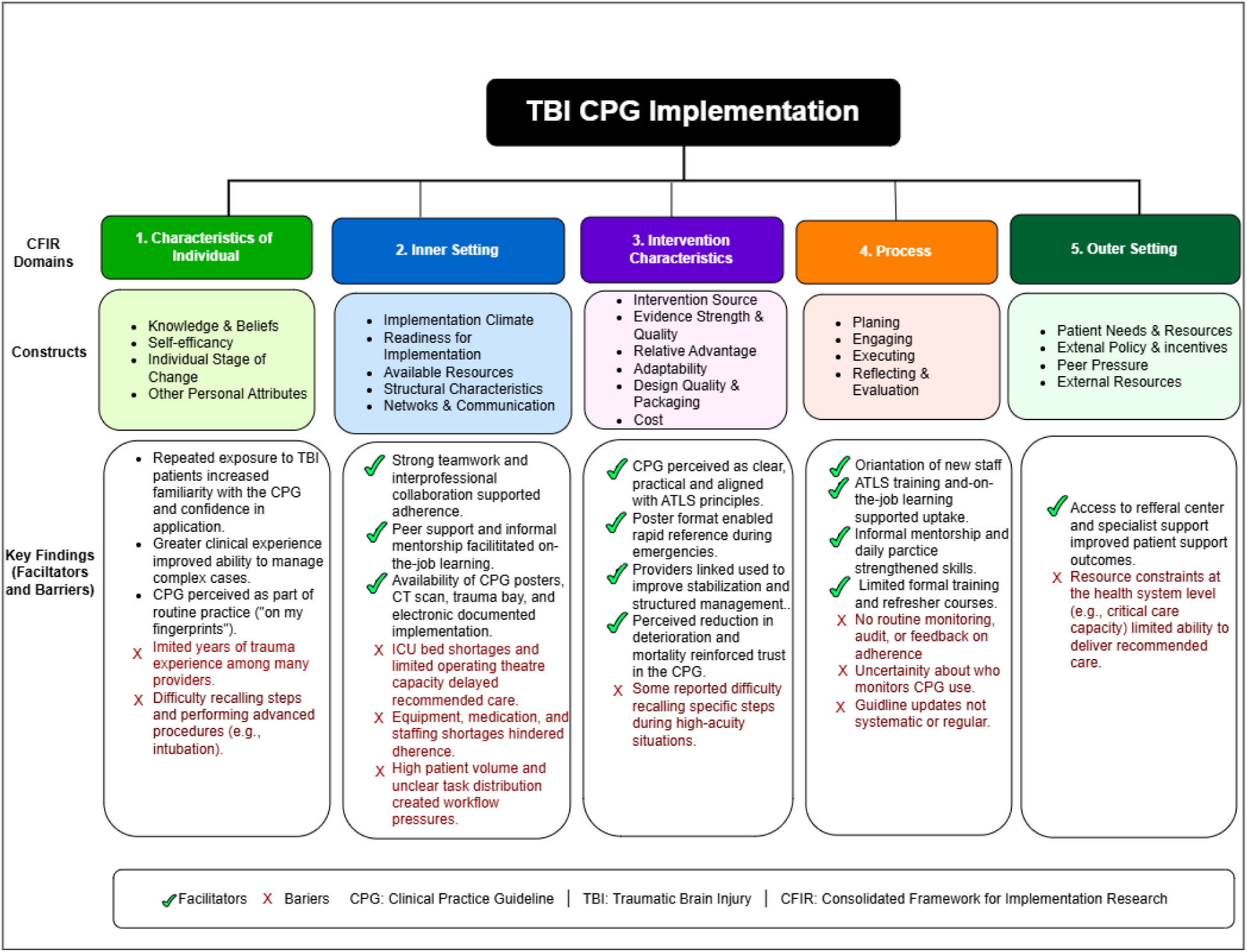
Healthcare Providers experiences facilitators, and barriers to implementation of the TBI CPG mapped to the CFIR domains and constructs

**Table 1:** Healthcare providers’ demographic characteristics (N=15)

| **Variable** | **Description, N (%)** |
| --- | --- |
| **Age:** mean (±SD) | 32 (±6.7%) |
| **Gender** |  |
| Male | 8 (53%) |
| Female | 7 (47%) |
| **Occupation** |  |
| Nurse | 7 (47%) |
| Doctors | 8 (53%) |
| **Doctors' Level of Education** |  |
| Specialist | 3 (20%) |
| Resident | 2 (13%) |
| MD | 2 (13%) |
| Intern doctor | 1 (7%) |
| **Nurses' Level of Education** |  |
| Nurse Specialist | 3 (20%) |
| Registered Nurse | 2 (13%) |
| Enrolled Nurse | 1 (7%) |
| Intern Nurse | 1 (7%) |
| **Presence of HCPs** |  |
| Before CPG | 2 (13%) |
| After CPG | 13 (87%) |
| **Years of experience at KCMC: mean (±SD)** | 5 (±5.2) |
| **Years of experience at EMD: mean (±SD)** | 4 (±2.5) |

**Table 2.** Identified themes and subthemes: Providers’ perceptions of the barriers and facilitators of the implementation of Clinical Practice Guidelines for Acute Management of Traumatic Brain Injury as stratified by the Consolidated Framework for Implementation Research (CFIR).

| CFIR Domain | Study Theme | Sub-theme | Interpretation |
| --- | --- | --- | --- |
| Characteristics of Individuals | Healthcare providers' experiences | Clinical experience | Repeated exposure to TBI patients strengthened familiarity with the guideline and clinical decision-making. |
|  |  | Professional experience in EMD/Surgery/Anaesthesia | Years of experience influenced confidence and ability to implement the CPG. |
|  | Facilitators | Feeling confidence | Frequent use increased self-efficacy and helped providers internalise the guideline into routine practice. |
|  | Barriers | Lack of clinical competence in performing some procedures. | Some providers lacked confidence in complex procedures (e.g., intubation) and struggled to recall guideline steps during emergencies. |
| Inner Setting | Facilitators | Effective teamwork | Effective teamwork, communication, and collaborative practice supported CPG adherence. |
|  |  | Existing infrastructure | Availability of CT imaging, trauma bay, equipment, and organized workflows facilitated implementation. |
|  |  | Managerial and institutional support | Leadership support, reporting structures, and departmental reminders promoted implementation. |
|  | Barriers | Organisational Barriers | Shortages of ICU beds, ventilators, monitoring equipment, medications, and operating theatre access limited adherence. |
|  |  | Workforce and workload challenges | Staff shortages and high patient volumes reduced the ability to complete all recommended CPG steps. |
| Intervention Characteristics | Facilitators | Positive experiences with CPG use | Perceived improvements in patient outcomes, safety, and systematic care increased trust in the guideline. |
|  |  | Structured and accessible guideline | Clear, evidence-based content and visual posters made the CPG easy to follow during emergencies. |
| Process | Facilitators | Orientation to the CPG | Initial departmental orientation introduced new staff to the guideline. |
|  |  | On-the-job learning and peer mentorship | Informal supervision and repeated practice reinforced implementation. |
|  |  | Prior trauma training (e.g., ATLS) | Previous evidence-based trauma education facilitated CPG uptake. |
|  | Barriers | Limited formal training and refresher sessions | Absence of structured continuing education reduced consistent implementation. |
|  |  | Self-directed learning | Some providers depended on independently reading the guideline rather than formal instruction. |
|  |  | Limited monitoring, evaluation, and guideline updating | Lack of audit, feedback, and regular revision weakened long-term implementation. |
| Outer Setting |  | None identified | Participants' accounts did not identify a dominant external policy or patient-level influences. |

### Characteristics of Individuals

#### Clinical experience and confidence

Clinical experience emerged as an important theme shaping health care providers’ perceptions of TBI CPG implementation. Participants described their experience by the number of TBI patients they treated and their years of experience in EMD, surgery, or anesthesia. They perceived that repeated exposure to TBI patients increased their familiarity with guidelines and their confidence in applying them.

> *“About a thousand patients over the three and a half years (INT 001).*
>
> *“In the two years I’ve been at KCMC, there are days when I treat around five patients, and there are days when I treat two. So, in a week, you can have more than ten TBI patients.” (INT 014)*

Participants generally expressed confidence in using guidelines, though confidence varied with clinical experience. Some described the CPG as part of their daily practice, whereas others reported difficulty recalling guideline steps or performing advanced procedures such as intubation.

> *“I am confident; I would say 100%.” (INT 012)*
>
> *“I have no records of that, yeah, because the CPG now is on my fingertips on a day-to-day basis even if I don’t refer” (INT 013).*

Participants described limited competence in performing certain recommended procedures as a barrier to applying the TBI CPG protocol, particularly intubation.

> *“I am not very comfortable with the intubation part. I had a severe TBI patient, and securing the airway is a challenge for me. The intubation part is kind of tough for me, so I solved it by looking for an alternative by asking for help because it’s teamwork” (INT 003).*

### Interventional characteristics

#### Structured and Accessible Guideline

Participants highlighted the TBI CPG as clear, practical, and aligned with international standards, including Advanced Life Support (e.g., ATLS). CPG posters in the trauma bay enabled rapid reference during emergencies and supported standardised patient assessment. Many participants also described positive experiences after using the guideline, reporting improved patient stabilisation and more structured clinical management.

“Participants suggested that the TBI CPG that is being used here at KCMC has a similar protocol as the Advanced Trauma Life Support strategy, and both are useful in managing TBI patients” *(INT 013)*.

> *“There is a board in the trauma area where you would see how to assess a patient suspected of having TBI.” (INT 006)*
>
> *“When you use it and see that your patient is not deteriorating” (INT 004)*

### Inner setting

#### Effective teamwork

Participants described that working as a team enables them to support one another during TBI management, making it easier to follow the guideline. They explained that colleagues frequently reminded one another of the guideline steps and supported decision-making during emergency care. Shared responsibility helped maintain continuity of patient management even when workload was high.

> *“First, we work as a team…once you start attending to a patient, it becomes easy to follow the guideline because someone will instruct you,’We start with this, then that…’ and it is something new every day, so we become aware.” (INT 016)*

#### Existing Infrastructure

Participants reported that organisational resources, including CT availability, a designated trauma bay, and electronic documentation systems, supported implementation of TBI CPG. These resources enabled more systematic assessment and management of patients with TBI.

> *“Now that we have a CT scan…patients come in, they are given oxygen, and they are intubated because there are people who are conversant with the protocol.” (INT 012)*

#### Managerial and Institutional Support

Participants described managerial and institutional support as an important facilitator of CPG implementation in ED. They reported that support from department heads and nurse coordinators, along with reporting mechanisms, encouraged adherence to the guideline and reinforced its use in routine clinical practice.

> *“All necessary investigations are available. We have equipment; we have two ventilators” (INT 007).*
>
> *“Based on the current system, we first have to report to the heads of departments, then they will take them forward to the responsible decision makers.” (INT 016)*
>
> *“For effective utilisation of the guideline…we should have at least weekly reminders on different protocols at the department.” (INT 016)*

#### Organizational barriers

Participants consistently described resource limitations as major barriers to CPG implementation. Shortages of ICU beds, operating room capacity, monitoring equipment, medications, and staffing frequently delayed recommended interventions. High patient volumes further limited providers’ ability to complete the guideline steps, particularly during busy shifts.

> *“The CPG recommends ICU admission, but sometimes the ICU is full.” (INT 013)*
>
> *“The workload is high compared to the number of staff…sometimes you cannot do all the CPG steps.” (INT 014)*
>
> *“It takes time before neurosurgery comes, and sometimes patients deteriorate before review.” (INT 007)*

### Process

Participants described implementation as largely supported by informal orientation, peer mentorship, and routine learning during clinical practice. While these mechanisms helped HCPs become familiar with the TBI CPG, many participants reported a lack of formal training, refresher sessions, routine audits, and regular CPG updates. Several participants also expressed uncertainty about who monitored adherence to the CPG.

> *“Ever since I attended here, I never received any training…just informal learning from exposure.” (INT 013)*
>
> *“The CPG was developed about 5–8 years ago, so it needs to be updated.” (INT 011)*
>
> *“I don’t know if there is a follow-up to check whether we use the CPG or not.” (INT 012)*

Participants described several gaps in implementing the TBI CPG. Formal orientation and refresher training were limited, and many providers reported learning about the guideline primarily through self-directed learning and routine clinical practice. Participants explained that new staff were often expected to familiarise themselves with the guideline independently, with additional guidance obtained from colleagues when needed. Although this approach helped providers become familiar with the CPG, participants reported limited opportunities for structured training and periodic reinforcement. In addition, some participants expressed uncertainty about whether the CPG was regularly reviewed or updated, raising concerns about its continued alignment with current clinical practice.

> *“The first time I was in the department, we had a patient who had severe TBI, and the specialist on call told us there is a poster on the notice board that we have to review and go through it” (INT 004).*
>
> *“Regarding the guideline, we train one another at the trauma bay, and this occasionally happens at least once a month. When they notice that we have a lot of new faces in the department, then we get a training” (INT 007).*
>
> ***“****No formal training.” “No updates of CPG, only that I know” (interview 006)*

### Outer setting

Participants did not identify substantial external policy or regulatory influences on implementation; instead, they primarily described organizational and health system resource constraints on CPG implementation in ED.

## DISCUSSION

Using the Consolidated Framework for Implementation Research (CFIR) allows us to explore healthcare providers’ experiences with the TBI Clinical Practice Guideline (CPG), identify key facilitators, and reduce barriers to CPG implementation. Findings show that although the guideline is generally accepted and integrated into routine practice, implementation remains shaped by provider experience, organisational support, and systemic resource constraints.

Providers perceived clinical experience as an important influence on TBI CPG implementation. Providers who managed high volumes of TBI cases reported greater familiarity with, and confidence in, internalizing the guideline and applying its recommendations. Repeated clinical exposure appeared to transform the CPG from an external reference into an internalised framework and a routine cognitive framework (“on my fingertips”), indicating a progression from external guidance to embedded clinical reasoning within the CFIR characteristics of the individual’s domain. This aligns with experiential learning theory and the implementation literature, which show that repeated protocol use strengthens adherence and skill acquisition, particularly knowledge and self-efficacy. Previous TBI research similarly indicates that clinicians’ professional experience, clinical context, and confidence influence the transition of evidence and guideline recommendations into practice (22–27).

Despite high patient volumes, many providers had relatively limited experience in emergency or trauma care. Participants also described challenges recalling guideline steps during emergencies and a lack of confidence in performing advanced procedures such as endotracheal intubation. These findings suggest that although clinical experience contributes substantially to competence, structured education and formal training may not fully replace it. Strengthening mentorship and competence development may therefore improve providers’ confidence and consistency in applying the guideline.

The physical environment also supported the implementation. Participants described the availability of posters in the trauma bay, CT imaging, and designated trauma space as strong facilitators of clinical decision-making and comprehensive patient assessment (28,29). Visible reminders can improve compliance with clinical guidelines, especially in acute care settings (30). Within CFIR, these findings align with the inner setting domain, particularly implementation climate and available resources. Access to imaging and specialized personnel further enhanced providers’ ability to deliver guideline-recommended interventions.

Interprofessional collaboration was a critical facilitator in TBI CPG implementation. Participants highlighted strong teamwork and effective communication (CFIR – Inner Setting) as key to sustaining implementation. Collaborative practice enables the providers to guide one another through guidelines and reinforces shared responsibility for patient care. Similar findings have been reported in trauma and emergency settings, where positive teamwork and interprofessional collaboration improve adherence to guidelines (31,32).

Despite these strengths, participants reported organizational barriers that limited adherence to the guideline. Resource limitations, including ICU bed shortages, limited operating theatre capacity, inadequate monitoring equipment, and medication stock-outs, directly interfered with timely management. High patient volume combined with limited staffing constrained providers’ ability to complete all guideline-recommended steps (33). These findings align with evidence from low-and middle-income countries (LMICs), where infrastructure limitations remain a major barrier to implementing trauma guidelines (33–36). Within CFIR, these challenges reflect the inner setting (structural characteristics). Importantly, these barriers did not reflect resistance to the guideline itself but rather contextual limitations that prevented full implementation.

Participants perceived the TBI CPG as clear, practical, and aligned with internationally recognised trauma management. The guideline’s poster format facilitated rapid reference during management and supported standardized patient assessment. Providers linked guideline use to reduced deterioration, structured management, and perceived declines in mortality (37–40). These positive clinical feedback loops reinforced trust in the CPG and strengthened motivation to continue using it (44, 45).

Within the CFIR, these findings reflect the intervention characteristics of evidence strength and quality, relative advantage, and design quality. When providers directly link a guideline to improved patient outcomes, adherence becomes intrinsically motivated rather than externally enforced. Confidence in and internalization of the CPG into routine practice further indicates deep integration into workflow, reflecting progression from initial adoption to normalization (43,44).

New-staff orientation, on-the-job training, and informal mentorship from experienced personnel supported implementation. Participants explained that they learned the guideline through routine practice, peer support, and some prior trauma training, such as Advanced Trauma Life Support (ATLS). These implementation activities promoted familiarity with the guideline despite limited formal education.

Nevertheless, participants identified important weaknesses in the implementation process. Formal orientation, refresher training, audit, and feedback mechanisms were inconsistently available, and many providers relied on self-directed learning after joining the department.

Participants also noted that the guideline had not been regularly reviewed or updated. These findings correspond to the CFIR process of engaging, reflecting, and evaluating, emphasizing that successful implementation requires continuous education, monitoring, and iterative improvement. Previous implementation research similarly shows that audit and feedback, ongoing training, and structured implementation support are essential for sustaining adherence to clinical practice guidelines (45,46).

Although participants discussed external issues related to CPGs, such as policies and patients’ needs, less frequently, health system limitations affected implementation of the TBI CPG. Limited access to critical care services, specialist services, and hospital capacity for TBI patients influenced providers’ ability to deliver guideline-recommended care. These contextual constraints show that implementation of evidence-based interventions in LMICs is influenced not only by provider behavior but also by broader health system capacity. Addressing these system-level challenges will be important for improving adherence to TBI guidelines and achieving optimal patient outcomes.

### Implications for Practice, Policy, and Research

The findings indicate that the TBI CPG has been well received and increasingly internalised among experienced providers, reflecting strengths in the Characteristics of Individuals domain, particularly knowledge, self-efficacy, and experiential learning from repeated exposure to TBI cases. Participants identified facilitators such as teamwork, access to visible guidelines in the trauma bay, and informal mentorship, underscoring the importance of a supportive inner setting, including a collaborative culture and an implementation climate.

However, sustaining CPG adherence requires strengthening structural and organizational mechanisms beyond individual commitment. Providers emphasized the need for structured orientation for newly recruited staff, periodic refresher training, and formal audit and feedback systems to reinforce CPG adherence. Persistent outer and inner setting constraints, including ICU bed shortages, limited equipment availability, and high workload pressures, were perceived as major barriers to guideline adherence, despite positive attitudes toward its use. Addressing these structural limitations while institutionalizing mentorship and continuous capacity building may enhance sustainability and ensure consistent, system-level adherence to TBI guideline-based care.

From a policy perspective, the findings underscore the need for health system investments that support guideline implementation, including adequate staffing, availability of ICUs for neurological patients, including those with TBIs, essential equipment, and integrating TBI guideline training into routine workforce programmes. Policymakers should consider establishing monitoring and evaluation mechanisms, such as audit-and-feedback systems, to promote accountability and sustained adherence to the TBI CPG.

From a research perspective, future studies should evaluate the effectiveness of implementation strategies, including structured guideline orientation programmes, mentorship, and audit-and-feedback interventions, in improving long-term CPG adherence and patient outcomes. Multi-site studies across diverse LMIC settings are also needed to assess the transferability of these findings and to identify context-specific factors that influence successful guideline implementation.

## Conclusion

This study shows that the TBI clinical practice guideline is well accepted and increasingly integrated into routine care, particularly among experienced providers, with strong teamwork and easy access to the guideline. However, structural limitations constrain sustained adherence, including resource shortages, workload pressures, and gaps in formal training and monitoring systems. Strengthening institutional support, continuous capacity building, and addressing critical infrastructure gaps are essential to ensure the sustainable use of TBI CPGs at KCMC EMD and in other LMIC settings.

## Authors Contribution

Vivian Frank Saria (PhD candidate and project principal investigator): conceptualization, study design, investigation, data collection, analysis, data interpretation, manuscript drafting, critical review and editing, project administration, and accountability for accuracy and integrity of all aspects of the work.

Gregory G. Zaccheus: Conceptualization, data collection, formal analysis, writing, review & editing

Blandina T. Mmbaga: Conceptualization, supervision, methodology, manuscript writing support, manuscript review

Catherine A. Staton: Conceptualisation, supervision, methodology, trauma and injury expertise, interpretation of findings, manuscript review and editing.

Anna Tupetz: methodology, data interpretation, formal analysis, agreement procedures, manuscript review and editing

Joao Ricardo Nickenig Vissoci: Conceptualization, methodology, manuscript review Francis Sakita, MD: Conceptualization, methodology, manuscript review

Winfrida C. Mwita-methodology, manuscript review and editing

Linda Minja: conceptualization, methodology advice, and manuscript review João Vítor Perez de Souza: Conceptualization, methodology, manuscript review. Kim Madundo: Conceptualization, manuscript review

## Institutional Review Board Statement

This study protocol was approved by the KCMC University Research and Ethics Committee (KUREC), IRB number 2545.

## Informed Consent Statement

All participants provided written or verbal informed consent in Kiswahili.

## Data Availability Statement

The data that support the findings of this study are available from the corresponding author, C.A.S., upon reasonable request.

## Data Availability

The qualitative data underlying the findings of this study are not publicly available due to participant confidentiality considerations. De-identified data are available from the corresponding author upon reasonable request.

## Acknowledgements

We thank Kilimanjaro Christian Medical Center (KCMC), KCMC University, and the staff of the Emergency Medicine, General Surgery, and Anaesthesia departments for their institutional and logistical support. We thank all healthcare providers who participated in the study and shared their experiences.

